# Association of mitochondrial DNA copy number with cardiometabolic diseases in a large cross-sectional study of multiple ancestries

**DOI:** 10.1101/2020.04.20.20016337

**Authors:** Xue Liu, Ryan J. Longchamps, Kerri Wiggins, Laura M. Raffield, Lawrence F. Bielak, Wei Zhao, Achilleas Pitsillides, Thomas Blackwell, Jie Yao, Xiuqing Guo, Nuzulul Kurniansyah, Bharat Thyagarajan, Nathan Pankratz, Stephen S. Rich, Kent D. Taylor, Patricia A. Peyser, Susan R. Heckbert, Sudha Seshadri, L Adrienne Cupples, Eric Boerwinkle, Megan L. Grove, Nicholas Larson, Jennifer A. Smith, Ramachandran S Vasan, Tamar Sofer, Annette L Fitzpatrick, Myriam Fornage, Jun Ding, Adolfo Correa, Goncalo Abecasis, Bruce M. Psaty, James G. Wilson, Daniel Levy, Jerome I. Rotter, Joshua C. Bis, Claudia L Satizabal, Dan E. Arking, Chunyu Liu

## Abstract

**Aims:** We tested the hypothesis that mitochondrial DNA copy number (CN) is associated with cardiometabolic disease (CMD) traits.

**Methods and results:** We determined the cross-sectional association of mtDNA CN measured in whole blood with several CMD traits in 65,996 individuals (mean age 60, 54% women, and 79% European descent). Cohort- and ancestry/ethnicity-specific association analysis was performed adjusting for trait- and cohort-specific covariates. Age was slightly positively associated with age (0.03 s.d. / 10 years (95% CI=0.01, 0.05)) before 65 years, while every 10 years older age was associated with 0.14 s.d. lower level of mtDNA CN after 65 years (95% CI= -0.18, -0.10). In meta-analysis without adjustment for white blood cell (WBC) and differential count in participants of European descent (N=52,491), low mtDNA CN was associated with increased odds of obesity (OR with 95% CI=1.13 (1.11, 1.16), P=3.3e-30) and hypertension (OR=1.05 (1.03, 1.08), P=4.0e-07). Further adjusting for WBC and differential count in the same participants of European descent (N=44,035), associations became non-significant (P>0.05) for hypertension, attenuated for obesity (OR_without cell count_=1.15 (1.12, 1.18) versus OR_cell count_=1.06 (1.03, 1.08)) but strengthened for hyperlipidemia (OR_without cell counts_ =1.03 (1.00, 1.06) versus OR_cell counts_ =1.06 (1.03, 1.09)). The magnitude and directionality of most associations were consistent between participants of European descent and other ethnicity/ancestry origins.

**Conclusion:** Low levels of mtDNA CN in peripheral blood were associated with an increased risk of CMD diseases.

## Introduction

Mitochondria convert dietary calories to molecular energy through oxidative phosphorylation (OXPHOS).^1^ In addition, mitochondria play essential roles in cellular differentiation, proliferation, reprogramming, and aging.^2-7^ Mitochondria contain their own genome (mtDNA) which is a circular, double-stranded DNA molecule of 16.6 kb. mtDNA encodes 13 key OXPHOS proteins, 22 transfer RNAs (tRNAs), and two ribosomal RNAs (rRNAs)^1^. Multiple copies of mtDNA are present per mitochondrion, and cells contain up to 7000 mitochondria per cell.^8^ The mtDNA copy number (mtDNA CN) correlates with cellular ATP generating capacity and metabolic status,^9^ and therefore, varies greatly across tissue and cell types depending on cellular energy demand.^1,10,11^

Several previous studies have demonstrated that mtDNA CN is lower in older individuals and this decrease is associated with a general decline in health.^12-14^ Low mtDNA content was also associated with higher fasting blood glucose (FBG), hemoglobin A1c (HbA1c), and lipid levels including high density lipoprotein (HDL), low density lipoprotein (LDL), triglycerides (TRIG), and total cholesterol (TC) in both diabetic patients and controls.^15^ In addition, mtDNA content was inversely related to BMI and fat accumulation in 94 healthy young individuals (mean age 30 years).^16^ A more recent study in two independent cohorts of women of European origin (n=2,278, mean age 30 years; and n=2,872, mean age 69 years), however, failed to detect significant associations between mtDNA CN and CMD risk factors including systolic blood pressure (SBP), diastolic blood pressure (DBP), HDL, LDL, TRIG, and glucose levels.^17^

Given inconsistent findings in previous studies and the central role of mtDNA in metabolism, we set out to investigate the association between mtDNA CN and several CMD risk factors in eight US cohorts representing participants of different ethnicity/ancestry origins with whole genome sequencing (WGS) and extensive cardiometabolic phenotyping. We also included individuals with Whole Exome Sequencing (WES) from the UK Biobank for validation. We performed cohort- and ancestry-specific association analysis between mtDNA CN and several CMD phenotypes. We also performed meta-analyses separately in participants of European descent and African Americans, and also combining all ancestry groups.

## Methods

### Study participants

Several cohorts from the NHLBI’s Trans-Omics for Precision Medicine (TOPMed) program contained a small number of duplicated participants. We removed one copy of the duplicated participants and this study included 26,890 individuals with WGS in the TOPMed program (67.4% women; age range of 20-100 years; 45.4% European Americans, 32.6% African Americans, 19.6% Hispanic/Latino Americans and 2.4% Chinese Americans) (**Supplemental Table 1**). Additionally, we included 39,106 participants of White British from the UK Biobank with WES (54% women; 40-75 years) for validation (**Supplemental Table 1**). All study participants provided written informed consent for genetic studies. The protocols for WGS and WES were approved by the institutional review boards (IRB) of the participating institutions (**Supplemental Materials**).

### mtDNA copy number estimation

*mtDNA CN estimation in WGS*: whole blood derived DNA was used for WGS through TOPMed sequencing centers. The average coverage was ~39x across samples. The program *fastMitoCalc* of the software package *mitoAnalyzer* was used to estimate mtDNA copy number across TOPMed participants.^13^ The average mtDNA CN per cell was estimated as twice the ratio of average coverage of mtDNA to average coverage of the nuclear DNA (nDNA). The coverage was defined as the number of reads that were mapped to a given nucleotide in the reconstructed sequence.^13^

*mtDNA CN estimation in UK BioBank:* whole blood derived DNA was used for WES in UK BioBank. mtDNA CN estimates were generated by customized regression with specific terms in Perl and R software using Exome SPB CRAM files (version Jul 2018) downloaded from the UK BioBank data repository (**Supplemental Materials**).

*mtDNA CN estimation in ARIC using other methods:* mtDNA CN estimation from low-pass WGS was calculated as the ratio of mitochondrial reads to the number of total aligned reads (**Supplemental Materials**). mtDNA CN estimated from the Affymetrix Genome-Wide Human SNP Array 6.0 was calculated using Genvisis 15 software package (**Supplemental Materials**). The participants whose mtDNA CN were estimated from low-pass WGS and Affymetrix Genome-Wide Human SNP Array 6.0 were independent to each other and were independent to those with TOPMed WGS in the ARIC cohort, as previously described^18^ (**Supplemental Materials**).

### Cardiometabolic disease phenotypes

Metabolic disease phenotypes were mapped to the examinations when blood was drawn for DNA extraction for mtDNA CN estimates. Our primary analysis focused on four CMD phenotypes, obesity, hypertension (HTN), type 2 diabetes (T2D), and hyperlipidemia. Obesity was defined as body mass index (BMI) ≥30 (kg/m^2^). T2D was defined as fasting blood glucose ≥126 mg/dL or currently receiving glucose-lowering or diabetes medication (s). Hypertension (HTN) was defined as systolic blood pressure (SBP) ≥140 mmHg, or diastolic blood pressure (DBP) ≥90 mmHg, or use of antihypertensive medication(s) for blood pressure control. Hyperlipidemia was defined as fasting total cholesterol (TC) ≥200 mg/dL or triglyceride (TRIG) ≥150 mg/dL, or use of any lipid-lowering medication.

We also analyzed the association of mtDNA CN with continuous cardiometabolic traits: BMI, SBP, DBP, FBG, HDL cholesterol, LDL cholesterol, and TRIG levels. In the analysis of FBG, we excluded individuals with diabetes, defined as glucose value ≥126 mg/dL and/or taking glucose-lowering or diabetes medications.^19^ SBP and DBP values (mmHg) were derived from the averages of two measurements. We added 15 mmHg and 10 mmHg to SBP and DBP, respectively, for individuals taking any BP lowering medications.^20^ The total cholesterol (TC) measurements were divided by 0.8 for individuals using lipid treatment medications.^21^ LDL (mg/dL) was calculated as (TC - HDL - TRIG/5) in individuals with TRIG <400 mg/dL using imputed TC values.^21^ In analyses of FBG and lipid levels, we excluded individuals whose fasting status was not established. TRIG, LDL and HDL values were log-transformed to approximate normality. Other continuous outcome variables were not transformed.

Metabolic syndrome is a collection of risk factors that increase the risk for cardiovascular disease (CVD).^22^ We analyzed the presence of metabolic syndrome variable in relation to mtDNA CN. An individual was classified as having metabolic syndrome (0/1) if he/she had three of the five following conditions: ^22^ 1) obesity – waist circumference >40 inches in men and >35 inches in women; 2) hyperglycemia – fasting glucose ≥100 mg/dL or currently receiving glucose-lowering or diabetes medication; 3) dyslipidemia – triglyceride ≥150 mg/dL or on lipid-lowering treatment; 4) dyslipidemia – High density lipoprotein cholesterol <40 mg/dL in men or <50 mg/dL in women or on lipid-lowering treatment; and 5) hypertension – 130 mmHg systolic or >85 mmHg diastolic or the current use of antihypertensive medication (s). Of note, the thresholds in defining metabolic syndrome are different from those for individual disease phenotypes in our primary analysis. Waist circumference was not measured in approximately a third of the FHS participants. Because BMI is the most common measure of overall obesity,^23^ to increase the sample size we used BMI ≥30 to define obesity in FHS participants with missing waist circumference values.

### Statistical analyses

In all analyses, we used mtDNA CN as the primary independent variable. To identify confounders and covariates in association analyses, we first examined whether mtDNA CN levels were associated with ‘blood collection year’ (the year when blood was drawn, as a surrogate of batch effects for blood-derived DNA samples) in all participating cohorts. White blood cell (WBC) count and blood differential count were previously reported to be associated with mtDNA CN.^18,24^ Therefore, we investigated whether mtDNA CN was associated with total WBC count, blood differential count, and platelet count in cohorts that measured or imputed^25,26^ these variables (**Supplemental Table 2**). We further examined mtDNA CN in relation to age and sex after adjusting for blood collection year (**Supplemental Figure 1**).

Based on observing significant associations of mtDNA CN in relation to ‘blood collection year’, age and sex, we generated mtDNA CN residuals for downstream analyses by regressing mtDNA CN on age, age squared, sex and blood collection year (as a factored variable) in each cohort. The residuals were standardized to a mean of zero and standard deviation (s.d.) of one, and used as the main predictor in all regression models. In the primary analysis, we used logistic regression (for unrelated individuals) and mixed effects logistic regression model (related individuals) to analyze binary outcomes (i.e., obesity, HTN) in relation to mtDNA CN residuals. Because age, sex and BMI are important confounders or covariates for cardiometabolic traits, we further adjusted for sex and age as covariates in the analysis of obesity, and adjusted for sex, age, age-squared (only for HTN) and BMI as covariates in the analysis of T2D, hyperlipidemia, and HTN. We used linear effects models to analyze continuous outcome variables, adjusting for the same set of covariates as for the respective binary outcomes. For cohorts with family structure, we accounted for maternal lineage as random effects in linear or logistic mixed models. A maternal lineage was defined to include a founder woman with all of her children, and all grandchildren from daughters of the founder woman.^27^

We performed the discovery meta-analysis in European American participants in TOPMed with fixed effects (P_Q_>0.01) or random effects (P_Q_≤0.01) inverse variance method and performed validation analyses using participants of White British in UK Biobank. We further compared meta-analysis results in participants of European descent to those from other racial/ethnic origins in TOPMed cohorts. Finally, we performed inverse-variance meta-analysis to combine results from TOPMed and UK Biobank. The primary results included associations of mtDNA CN with the four disease outcomes. We used p=0.01 for significance to account for multiple testing for primary results, and used p=0.05/9~0.006 for significance in analysis of continuous outcomes.

Measured and/or imputed WBC variables were available in a subset participants in TOPMed and in all participants in UK Biobank. We compared associations between mtDNA CN and individual outcomes in the same participants with and without WBC count, differential count and platelet count as additional covariates.

We further investigated whether sex or age modified the association between mtDNA CN and outcome variables, adjusting for the same set of covariates described in primary analyses. We included an interaction term between mtDNA CN and sex/age in association analyses. We also performed stratified analyses between mtDNA CN and CMD traits in participants less than 65 years old and participants 65 years or above (see **Supplemental methods** for details). The statistical software R (version 3.6.0) was used for all statistical analyses.

## Results

### Characteristics of Study Participants

The current study included 13,385 European Americans, 8,012 African Americans, 601 Chinese Americans, and 4,892 Hispanic/Latino Americans from the TOPMed program as well as 39,106 White British from the UK Biobank. On average, 55% of study participants were women, and the participants’ mean age was 60 years (range 20 to 100 year; **Supplemental Table 1**). We observed moderate to high heterogeneity in distributions of age, sex, and cardiometabolic phenotypes across cohorts and ancestries. For example, the age range was 20 to 100 years in the Framingham Heart Study (FHS) (mean age 60 years, 40% pariticipants≥65 years). In contrast, all participants in the Cardiovascular Health Study (CHS) were older than 65 year of age (mean age 74). HTN, obesity, T2D, and hyperlipidemia were more prevalent in African Americans than participants of other ethnic and racial groups (**Supplemental Table 1**)

### A threshold effect between age and mtDNA CN

Because low mtDNA CN was associated with older age and reported to be associated with increased cardiometabolic disease risk,^15-17,28^ we reported beta estimates as the change in an outcome variable in response to 1 s.d. lower mtDNA CN in all of analyses. We observed that, on average, age was associated with a slightly higher level of mtDNA (0.032 s.d. / 10 years (95% CI= 0.013, 0.052), P=0.0014) of mtDNA CN from 20s to 65 years. We observed a threshold effect of age on mtDNA CN, and every 10 years older age was associated with 0.14 s.d. lower level of mtDNA CN after 65 years (95% CI= -0.18, -0.10), P=1.82e-13) (**Figure 2**). The relationship between mtDNA CN and age appeared to be similar in men and women. Women had higher mtDNA CN than men (beta=0.23, 95% CI= (0.20, 0.26), P=7.4e-60) as noted previously.^13,24^ The threshold effect between age and mtDNA CN was slightly attenuated after adjusting for WBC counts (**Supplemental Figure 2**).

**Figure 1.**
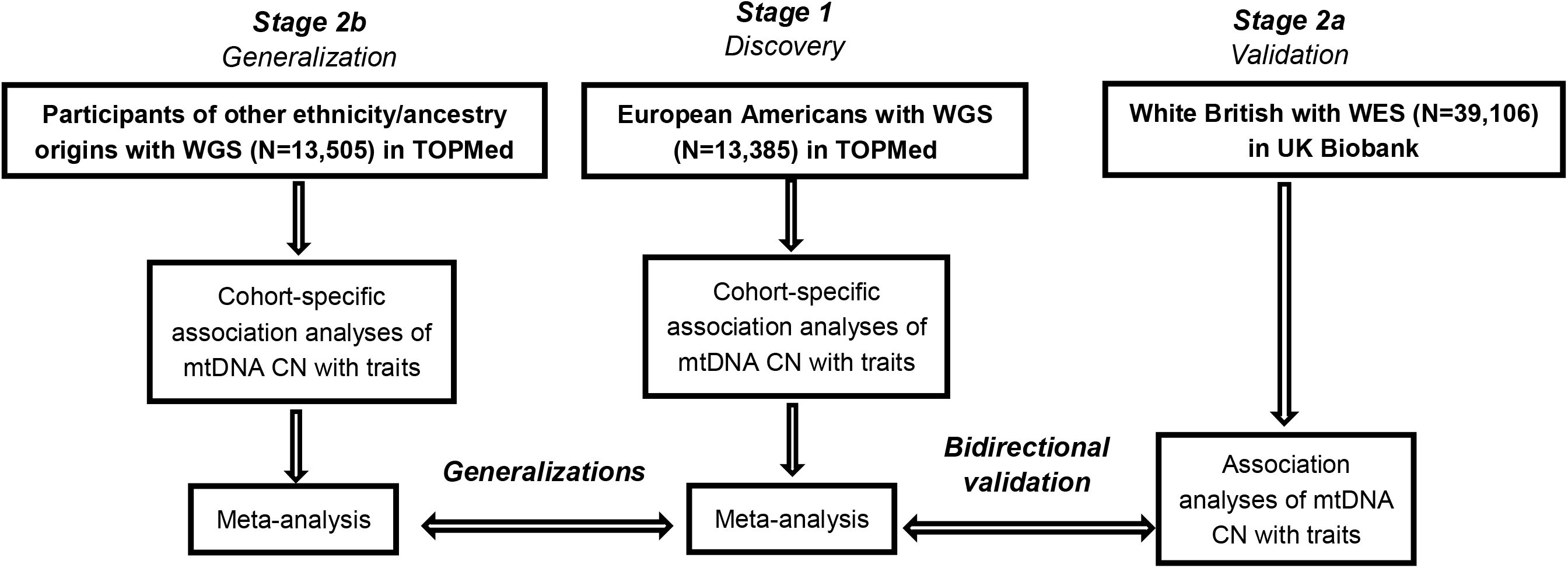
Study design. Association analysis of mtDNA CN with metabolic traits was performed in cohorts of European Americans (N=13,385), African Americans (N=8,012), Chinese Americans (N=601) and Hispanic and Latino Americans (N=4,892) in TOPMed and in White British participants of the UK Biobank (N=39,106). Meta-analysis using fixed or random effects inverse variance method was used to summarize the results in European Americans and African Americans in TOPMed. Primary analysis included age, age^2^ (blood pressure traits), sex, BMI (not in BMI traits) and batch effects. Second analysis included covariates in primary analysis and imputed/measured white blood cell count and blood differentials. Interaction analysis was performed to investigate whether age and sex modified the relationship between mtDNA CN and cardiometabolic disease traits.

**Figure 2.**
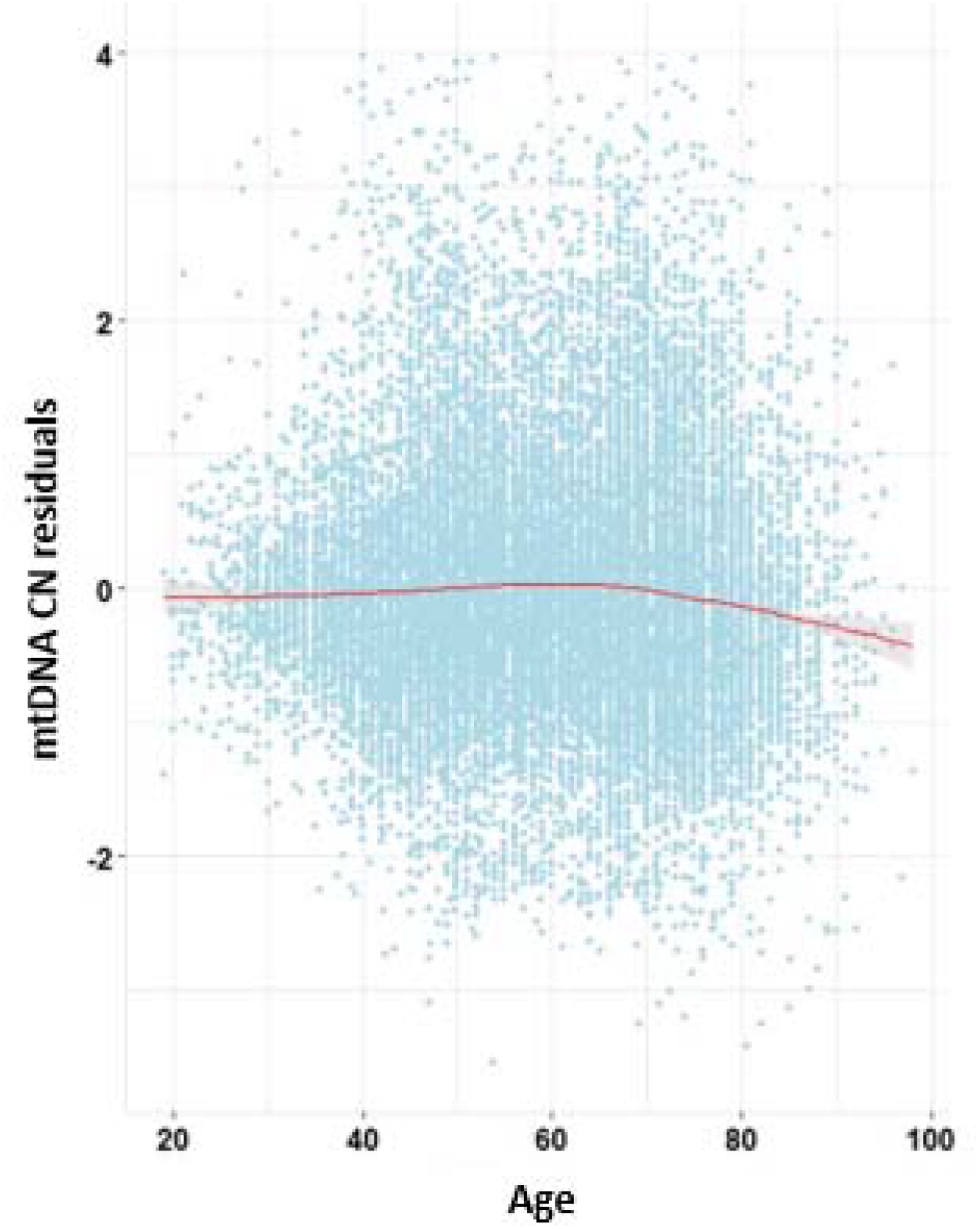
The relationship of mtDNA CN with age in TOPMed participants. Scatterplots of mtDNA CN residuals versus age. Cohort specific mtDNA CN residuals were obtained by regressing mtDNA CN on batch information.

### Discovery meta-analysis in European Americans participants

We performed the discovery meta-analysis in European American participants in TOPMed (N =13,385). We found that 1 s.d. decrease in mtDNA CN was significantly associated with 1.08-fold odds of obesity (95% CI= (1.03, 1.13), P=5.0e-04), 1.07-fold odds of hypertension (95% CI= (1.03, 1.12), P=1.4e-03), 1.16-fold odds of metabolic syndrome (95% CI= (1.06, 1.27), P=1.8e-03). We also found that 1 s.d. decrease in mtDNA CN was nominally (0.01<P<0.05) associated with 1.18-fold odds of diabetes (95%= (1.01, 1.37), P=0.041). For continuous traits, 1 s.d. decrease in mtDNA CN was significantly associated with 0.033 unit (95% CI= (0.023, 0.041), P=1.7e-04) increase in TRIG and nominally associated with 0.38 mmHg (95% CI= (0.021, 0.73), P=0.038) increase in SBP and 0.22 kg/m^2^ (95% CI= (0.074, 0.25), P=0.028) increase in BMI. mtDNA CN was not significantly associated with DBP, HDL, LDL, or fasting glucose (P>0.05) in the discovery meta-analysis (**Table 1, Figures 3 and 4**)

**Table 1.** Cross-sectional association and meta-analysis of mtDNA CN with metabolic disease phenotypes.

|  | Traits | Discovery (European Americans)<br>(N=13,385) |  |  | Replication (White British)<br>(N=39,106) |  |  | Meta-analysis<br>(N=52,491) |  |  |
| --- | --- | --- | --- | --- | --- | --- | --- | --- | --- | --- |
|  |  | OR | 95% CI | P-value | OR | 95% CI | P-value | OR | 95% CI | P-value |
| Binary outcomes | Obese | 1.08 | 1.03-1.13 | 5.0e-04 | 1.15 | 1.12-1.18 | 1.6e-30 | 1.13 | 1.11-1.16 | 3.3e-30 |
|  | HTN | 1.07 | 1.03-1.12 | 1.4e-03 | 1.04 | 1.02-1.07 | 0.00023 | 1.05 | 1.03-1.08 | 4.0e-07 |
|  | Diabetes | 1.18 | 1.01-1.37 | 0.041 | 1.04 | 0.99-1.10 | 0.10 | 1.05 | 1.00-1.10 | 0.051 |
|  | Hyperlipids | 1.04 | 0.99-1.08 | 0.11 | 1.03 | 1.00-1.06 | 0.024 | 1.03 | 1.00-1.05 | 0.023 |
|  | MetS | 1.16 | 1.06-1.27 | 1.8e-03 | 1.13 | 1.10-1.15 | 1.1e-24 | 1.13 | 1.11-1.16 | 9.7e-27 |
|  |  | Beta | SE | P-value | Beta | SE | P-value | Beta | SE | P-value |
| Continuous outcomes | BMI | 0.22* | 0.10* | 0.028* | 0.32 | 0.023 | 9.1e-42 | 0.31 | 0.023 | 2.5e-41 |
|  | DBP | 0.02* | 0.20* | 0.92* | 0.19 | 0.054 | 0.00038 | 0.15 | 0.049 | 1.9e-03 |
|  | SBP | 0.38 | 0.19 | 0.038 | 0.42 | 0.094 | 7.2e-06 | 0.42 | 0.085 | 6.0e-07 |
|  | FBG | 0.067 | 0.092 | 0.47 | 0.20 | 0.052 | 2.0e-04 | 0.16 | 0.046 | 5.4e-04 |
|  | HDL | -0.0097* | 0.0099* | 0.33* | -0.099 | 0.069 | 0.16 | -0.012 | 0.0098 | 0.21 |
|  | LDL | -0.008* | 0.0073* | 0.27* | -0.0040 | 0.0014 | 0.0045 | -0.0027 | 0.0014 | 0.059 |
|  | TRIG | 0.033 | 0.0088 | 1.7e-04 | 0.023 | 0.0025 | 8.6e-21 | 0.025 | 0.0022 | 8.3e-30 |

The beta estimates are in units of metabolic traits corresponding to one s.d. lower mtDNA-CN. Association analysis of mtDNA CN with metabolic traits was performed in cohorts of European Americans in TOPMed and also in White British participants in UK Biobank. Meta-analysis with fixed or random effects inverse variance method was used to summarize the results in European descent in TOPMed and White British in UK Biobank. DBP, diastolic blood pressure; SBP, systolic blood pressure; BMI, body mass index; FBG, fasting blood glucose; HDL, high density lipoprotein; Trig, triglyceride. Obese, obesity; HTN, hypertension; Diabetes, Diabetes; MetS, metabolic syndrome. Asterisk (\*) denotes inverse variance random effect meta-analysis.

**Figure 3.**
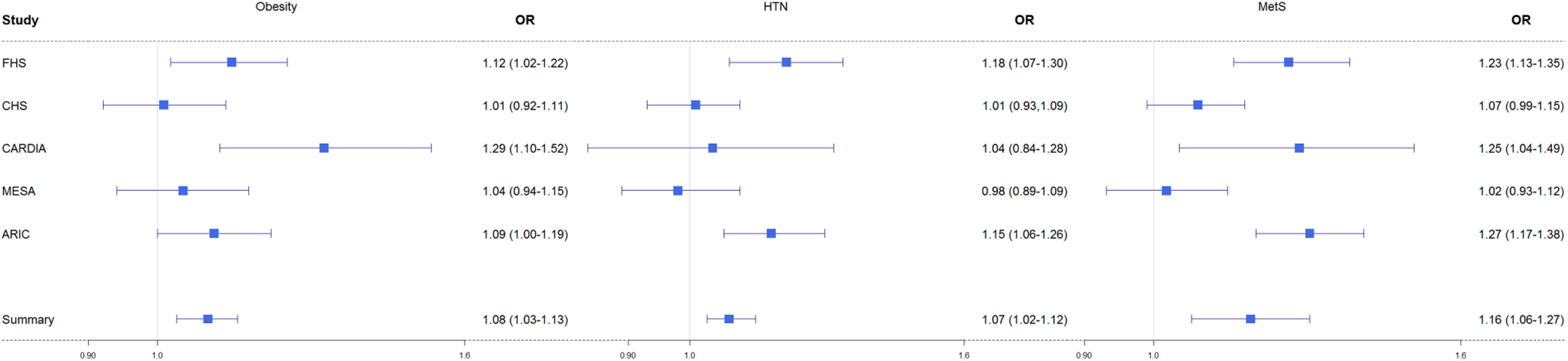
Association and meta-analyses of mtDNA CN with obesity and hypertension (HTN) and metabolic syndrome (MetS) in European Americans (N=13,385) in TOPMed. Fixed effects inverse variance method was used to summarize the results. The odds ratio (OR) corresponds to one s.d. decrease in mtDNA-CN. Covariates included age, sex, BMI (for HTN and MetS) and batch variables.

**Figure 4.**
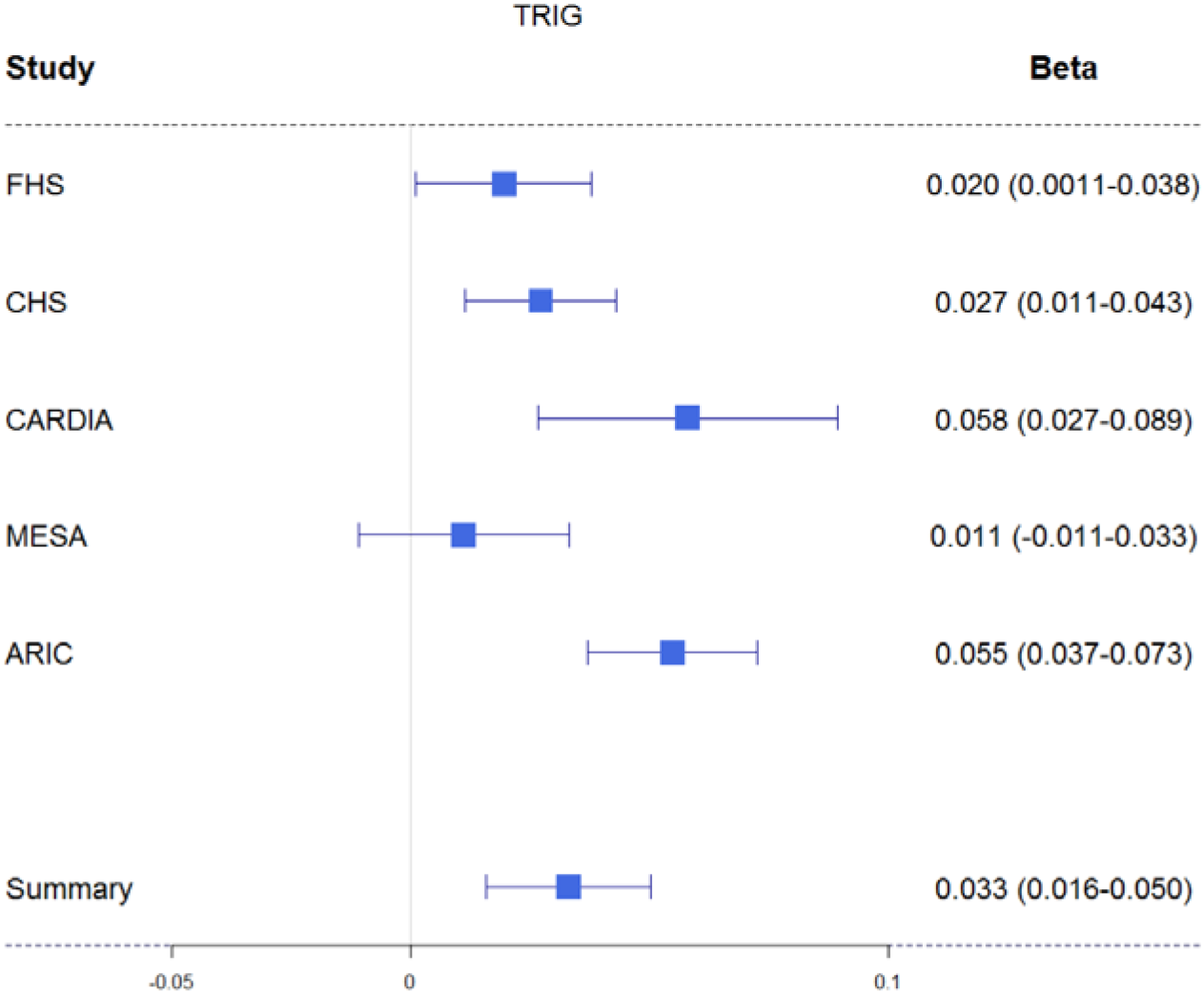
Association and meta-analyses of mtDNA CN with triglyceride in European Americans (N=13,385) in TOPMed. Fixed effects inverse variance method was used to summarize the results. The effect size estimates are in units of metabolic traits corresponding to one s.d. decrease in mtDNA-CN. TRIG, triglyceride. Covariates included age, sex, BMI (for HTN and MetS) and batch variables.

### Bidirectional validation and meta-analysis with participants of European descent between TOPMed and UK Biobank

Most of the significant associations in meta-analysis of the discovery phase were validated in the UK Biobank (**Table 1**). Compared to those in the discovery meta-analysis, the UK Biobank data yielded larger effect sizes for associations of several traits (**Supplemental Figure 3**). For example, 1 s.d. decrease in mtDNA CN was associated with 1.15-fold (95% CI= (1.12, 1.18)) odds of obesity in the UK Biobank versus 1.08-fold (95% CI= 1.03-1.13) in the TOPMed EA-specific meta-analysis. Additionally, 1 s.d. unit decrease in mtDNA CN was associated with 0.32 kg/m^2^ increase in BMI in UK Biobank (p = 9.1e-42) compared to 0.22 kg/m^2^ increase in BMI (p=0.028) in the meta-analysis of TOPMed EA participants. DBP and FBG were not significant in the discovery TOPMed EA meta-analysis (beta=0.02, 95% CI= (−0.37, 0.41), P= 0.92 for DBP; beta=0.067, 95% CI= (−0.11, 0.25), P=0.47 for FBG) while significant in UK Biobank (beta=0.19, 95% CI= (0.086, 0.30) with P=0.00038 for DBP; beta=0.20, 95% CI = (0.093, 0.30) with P=2.0e-4 for FBG) (**Table 1**).

We performed meta-analysis of all participants of European descent in TOPMed and UK Biobank (n=52,491) using fixed effects inverse-variance method to combine results (**Table 1**). One s.d. increase in mtDNA CN was significantly associated with an increase of odds of obesity (OR=1.13, 95% CI= (1.11, 1.16), P=3.3e-30), hypertension (OR=1.05, 95% CI= (1.03, 1.08), P=4.0e-07), metabolic syndrome (OR=1.13, 95% CI= (1.11, 1.16), P=9.7e-27), and nominally associated with diabetes (OR=1.05, 95% CI= (1.00, 1.10), P=0.051), hyperlipidemia (OR=1.03, 95% CI= (1.00, 1.05), P=0.023). For continuous phenotypes, 1 s.d. decrease in mtDNA CN was significantly associated with 0.15 mmHg increase in DBP (95% CI=(0.056, 0.25), P=1.9e-03), 0.42 mmHg increase in SBP (95% CI= (0.26, 0.59), P=6.0e-07), 0.32 kg/m^2^ increase in BMI (95% CI=(0.27, 0.36), P=2.5e-41), 0.16 mg/dL increase in fasting glucose (95% CI= (0.069, 0.25), P=5.4E-04), and 0.025 unit increase in TRIG (95% CI= (0.021, 0.029), P=8.3e-30).

mtDNA CN estimated from low-pass WGS and Affymetrix Genome-Wide Human SNP Array 6.0 gave rise to consistent associations for most of the CMD traits compared to that from WGS (**Supplemental Table 7**).

### Comparison of results from the European descent group with those from other racial/ethnic groups

The directionality of associations of mtDNA CN with CMD traits was consistent in African Americans (N=8,012), Hispanic/Latino Americans (N=4,892) and Asian Americans (N=601) compared to that in participants of European descent for most of the phenotypes (**Table 1**, **Figure 5** and **Supplemental Table 3, 4, Supplemental Figure 4, 5**). In the meta-analysis of AA participants, 1 s.d. decrease in mtDNA CN was significantly associated with 1.14-fold odds of diabetes (95%CI= (1.06, 1.22), P=2.5e-04) and a 0.68 mmHg increase in SBP (95% CI= (0.21, 1.15), P=4.0e-03). In Asian-only TOPMed participants (n=601), 1 s.d. decrease in mtDNA CN was significantly associated with 1.43-fold odds of hyperlipidemia (95% CI= (1.19, 1.72), P=0.00014). In Hispanic-only TOPMed participants (n=4,900), One s.d. decrease in mtDNA CN was significantly associated with an increase of odds of diabetes (OR=1.16, 95%CI= (1.06, 1.26), P=0.0012), and significantly associated with 0.019 unit decrease in LDL (95 % CI= (−0.029, -0.0079), P=6.6e-04) and 0.028 unit increase in TRIG (95% CI= (0.010, 0.046), P=0.0020). (**Supplemental Table 4**). Results of meta-analysis in pooled-samples are included in **Supplemental Table 5**.

**Figure 5.**
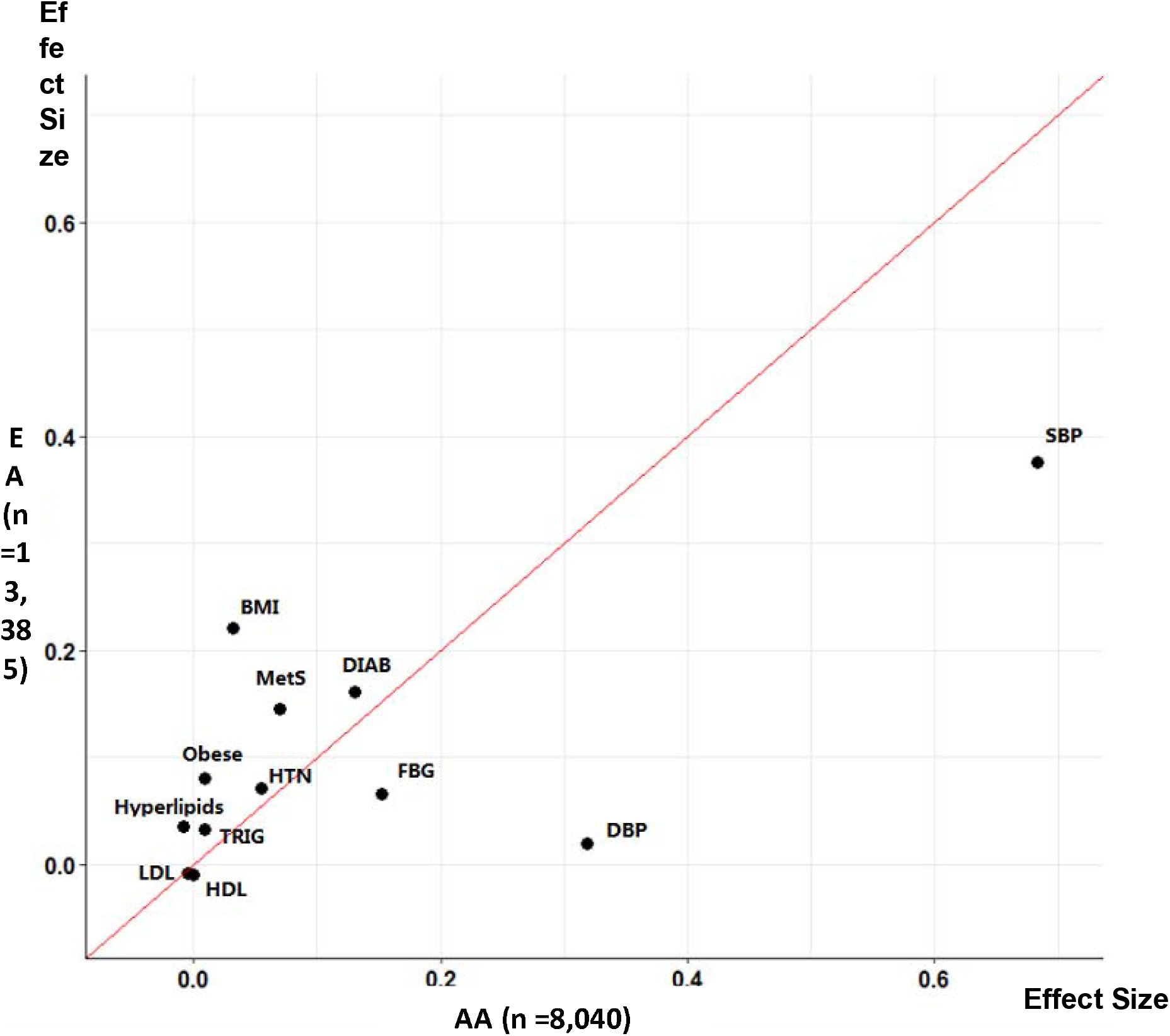
Comparison of beta of metabolic traits in the European Americans and American Americans in TOPMed. DBP, diastolic blood pressure; SBP, systolic blood pressure; BMI, body mass index; FBG, fasting blood glucose; HDL, high density lipoprotein; LDL, low density lipoprotein, TRIG, triglyceride. TC, total cholesterol; Obese, obesity; HTN, hypertension; Diabetes, Diabetes; Hyperlipids, hyperlipidemia; MetS, metabolic syndrome.

### Accounting for WBC and blood differential count as additional covariates

WBC count and blood differential count were available in a subset of participants in TOPMed (n=12,345) and in participants in the UK Biobank (n=39,021). mtDNA CN was inversely associated with the total WBC count and two blood cell differentials, neutrophil and monocyte. In contrast, mtDNA CN was positively associated with lymphocyte and platelet (**Supplemental Table 2**). Because the WBC count is associated with mtDNA CN and it is an indicator of a systemic subclinical inflammation state that accompanies CMD risk factors,^29-33 34-36^ we compared results between models with and without the total WBC count and blood differentials as additional covariates in the same participants. Directionality remained the same for all associations except for HDL after adjusting for WBC count and blood differential count in participants of European descent (**Supplemental Table 6, Supplemental Figure 6, 7**). Comparison of regression coefficients with and without adjusting for WBC count and differential in participants of other ethnicity and ancestry origins were available in **Supplemental Figure 8, 9**. Non-lipid CMD traits and TRIG attenuated their associations with mtDNA CN after adjusting for WBC cell counts and differentials. For example, the association of mtDNA CN with obesity was attenuated: OR_without cell counts_=1.15 (1.12, 1.18), P=7.7e-31 versus OR_cell counts_=1.06 (1.03, 1.08), P=2.5e-06. In contrast, hyperlipidemia and LDL were not significantly associated with mtDNA CN before adjusting for cell counts (P>0.05) and became significant after adjusting for cell counts and blood differential count (OR=1.06 (1.03, 1.09), P=3.9e-05 for hyperlipidemia and beta=0.012 (0.0096, 0.015), P=7.8e-18 for LDL) (**Supplemental Table 6**).

### Interaction analyses

We did not find statistically significant interactions between sex and mtDNA CN, or between age and mtDNA CN with respect to the phenotypes tested when including interaction terms in the models (**Supplemental Table 8**). Due to the threshold effect between age and mtDNA CN, we further performed association analyses in younger (<65 years) and older (≥65 years) participants. It appeared that the effect sizes were larger in younger individuals compared to older individuals for a number of traits although the directionality remained to be the same for these traits (**Supplemental Materials**, **Supplemental Table 9** and **Supplemental Figure 10**).

## Discussion

We demonstrated that low levels of mtDNA CN in peripheral blood were associated with an increased risk of CMD diseases in 65,996 individuals representing multiple ethnicity and ancestry origins. Consistent effect estimates were obtained from the discovery meta-analysis and the validation analysis in UK Biobank participants with adjustment for traditional clinical covariates as well as adjustment for WBC and blood differential count, demonstrating the robustness of our findings These findings further suggests that altered levels of mitochondrial energy production may be involved in the development of a cluster of conditions that increase the risk of CVD.

A longitudinal study reported that a 1 s.d. decrease in mtDNA CN was associated with 1.29-fold and 1.11-fold increased risk of developing CHD and stroke, respectively.^37^ Cardiometabolic factors are known risk factors for the development of CVD. This study, however, discovered only modest associations of mtDNA CN with several cardiometabolic (CMD) traits (odds ratio=1.04 to 1.16 across individual CMD traits and the composite metabolic syndrome), indicating that factors other than CMD may drive or mediate associations between mtDNA CN and cardiovascular outcomes. The mechanisms underlying mitochondrial dysfunction and CVD are largely unknown. One mechanism by which mtDNA CN may influence CVD is through the regulation of nuclear DNA methylation and gene expression. A mouse hybrid nuclear DNA and mtDNA system was previously developed, i.e., a hybrid mouse containing nuclear DNA from control mouse strain A and a specific mtDNA from control mouse strain b.^38,39^ Using this model, two studies provided direct evidence that different mtDNA background influences DNA methylation, gene expression, and cellular adaptive response.^38,39^ We recently assessed the relationship between mtDNA-CN and nuclear DNA methylation in up to 5,000 African American and European American study participants. Several validated mtDNA-associated CpGs were also known to be associated with coronary heart disease and CVD.^40^ Functional assessment demonstrated that modification of mtDNA CN levels resulted in changes in DNA methylation at specific CpGs and expression levels of nearby genes through knockout of the mitochondrial transcription factor, the protein that regulates mtDNA replication in CRISPR-Cas9.^40^ These results provided evidence that mtDNA CN may impact cardiovascular health through DNA methylation and differential expression of specific genes that may impact cell signaling. Further studies are needed to investigate whether DNA methylation and gene expression mediate the effects of mtDNA CN on cardiovascular risks.

WBC count is a blood biomarker of systemic inflammation. It has been increasingly recognized that chronic low-grade inflammatory state accompanies CMD risk.^29^ Previous studies reported that WBC count is associated with increasing risk of obesity,^30,31^ diabetes,^29,32,33^ hypertension,^34-36^ while heterogeneous relationships with lipid levels.^41^ This study and previous studies also showed that a high WBC count was associated with low mtDNA CN.^42-47^ Therefore, the connections between mtDNA CN, inflammation, and metabolic disease phenotypes are complex. These findings indicates that mtDNA CN and WBC count may interplay for CMD risk. Further studies are warranted to investigate underlying molecular mechanisms to establish a potential causal pathway among mtDNA CN, inflammation and CMD risk.

Most previous studies reported a tendency toward low mtDNA CN with advancing age,^12,13,18,24,48,49^ The current cross-sectional study, using a large sample size with a wide age range, demonstrated that mtDNA CN increases slightly from young adult (age 20 to 29) to late middle age (65 years old), and decreases in individuals older than 65 years. Most of the previous studies included participants with a limited age range, that is, the majority of study participants were young, middle-aged, or older individuals.^12,49,50^ In addition, post of these previous studies only investigated the linear relationship between mtDNA CN and age.^12,13,18^ Using a large sample size, this study discovered a threshold effect of age on mtDNA CN. However, this study only investigated age-mtDNA CN association cross-sectionally. The intra-individual mtDNA CN variability with advancing age needs to be explored in a longitudinal setting in future studies.

## Strengths and Limitations

This study includes a large sample size of men and women of multiple ethnicity and ancestry origins across a wide age range, thus enables us to investigate the relationships of mtDNA CN with CMD phenotypes throughout the entire adult life span. In addition, we performed careful phenotype harmonization and examined several potential confounding variables of mtDNA CN in association analysis with metabolic traits. Furthermore, we included association analyses of CMD traits with mtDNA CN estimated from several technologies further demonstrated the utility of mtDNA CN estimated from these different technologies in association analysis. Despite the multiple strengths in this study, several limitations should be noted. mtDNA CN was estimated using DNA derived from whole blood, which is not necessarily the relevant tissues with respect to cardiometabolic (e.g., cardiac muscle, skeletal muscle, adipose tissue) and aging-related (e.g., brain) disease phenotypes. Nevertheless, peripheral blood is easily accessible, changes in mtDNA in whole blood should reflect metabolic health across multiple systems. A recently study found that 23blood-derived mtDNA CN is associated with gene expression across multiple tissues and is predictive for incident neurodegenerative disease, which provides evidence supporting the hypothesis that changes in mtDNA in whole blood may reflect metabolic health across multiple systems.^51^ Second, though we accounted for confounders and known batch effects in mtDNA CN and harmonized metabolic traits, we still observed moderate to high heterogeneity in the association coefficients in meta-analysis of most of the phenotypes in both ancestry-specific analyses and across ancestries. Different distributions of age, sex, and phenotypes across study cohorts may partially explain the heterogeneity in these associations. Unobserved confounding factors, such as experiment conditions for blood drawing, DNA extraction, and storage may also have contributed to heterogeneity. Finally, we were not able to determine causal relationships between mtDNA CN and metabolic traits due to the cross-sectional nature of the study. Future studies with mtDNA CN and CMD traits at two time points will provide further insight into associations of aging-related mtDNA CN change with CMD traits.

## Data Availability

Raw data were generated at NHLBI's TOPMed program and can be requested upon approval from TOPMed. Derived data supporting the findings of this study are available from the corresponding author C.Liu on request.

## Acknowledgments

Detailed acknowledgment for each cohort is included in Supplemental Materials. In Brief, the authors thank the staff and participants of the Atherosclerosis Risk in Communities study, Cardiovascular Health Study, Coronary Artery Risk Development in Young Adults Study, Framingham Heart Study, Jackson Heart Study, Genetic Epidemiology Network of Arteriopathy Study, Hispanic Community Health Study/Study of Latinos, Multi-Ethnic Study of Atherosclerosis Study, and UK Biobank for their important contributions. We gratefully acknowledge the studies and participants who provided biological samples and data for TOPMed.Whole genome sequencing (WGS) for the Trans-Omics in Precision Medicine (TOPMed) program was supported by the National Heart, Lung and Blood Institute (NHLBI). Centralized read mapping and genotype calling, along with variant quality metrics and filtering were provided by the TOPMed Informatics Research Center (3R01HL-117626-02S1; contract HHSN268201800002I). Phenotype harmonization, data management, sample-identity QC, and general study coordination were provided by the TOPMed Data Coordinating Center (3R01HL-120393-02S1; contract HHSN2682018000011). Additional phenotype harmonization was performed by the current study (R01AG059727).

The views expressed in this manuscript are those of the authors and do not necessarily represent the views of the National Heart, Lung, and Blood Institute; the National Institutes of Health; or the U.S. Department of Health and Human Services.

## Translational Perspective

The mitochondrial genome (mtDNA) is represented at variable copy number (CN) in human cells and plays essential roles in cellular metabolism. We determined the cross-sectional association of mtDNA CN measured in whole blood with several cardiometabolic traits in 65,996 individuals (mean age 60, 54% women, and 79% participants of European descent). Low mtDNA CN levels were significantly associated with an increased risk of obesity and hyperlipidemia after accounting for clinical covariates and blood cell counts. The magnitude and directionality of associations were consistent between participants of European descent and other ancestries/ethnicities. Understanding the role of mtDNA CN in cardiometabolic will provide insight into the pathobiology underlying cardiometabolic diseases.

## References

1. Voet DVJ, Pratt CW. Fundamentals of Biochemistry. 2nd Edition. John Wiley and Sons, Inc. 2005:pp. 547. ISBN 0471214957..

2. Osellame LD, Blacker TS, Duchen MR. Cellular and molecular mechanisms of mitochondrial function. Best Pract Res Clin Endocrinol Metab 2012;26:711–23.

3. Antico Arciuch VG, Elguero ME, Poderoso JJ, Carreras MC. Mitochondrial regulation of cell cycle and proliferation. Antioxid Redox Signal 2012;16:1150–80.

4. Takahashi K, Yamanaka S. Induction of pluripotent stem cells from mouse embryonic and adult fibroblast cultures by defined factors. Cell 2006;126:663–76.

5. Misko AL, Sasaki Y, Tuck E, Milbrandt J, Baloh RH. Mitofusin2 mutations disrupt axonal mitochondrial positioning and promote axon degeneration. J Neurosci 2012;32:4145–55.

6. Clayton DA, Doda JN, Friedberg EC. The absence of a pyrimidine dimer repair mechanism in mammalian mitochondria. Proc Natl Acad Sci U S A 1974;71:2777–81.

7. Seo AY, Joseph AM, Dutta D, Hwang JC, Aris JP, Leeuwenburgh C. New insights into the role of mitochondria in aging: mitochondrial dynamics and more. J Cell Sci 2010;123:2533–42.

8. Miller FJ, Rosenfeldt FL, Zhang C, Linnane AW, Nagley P. Precise determination of mitochondrial DNA copy number in human skeletal and cardiac muscle by a PCR-based assay: lack of change of copy number with age. Nucleic Acids Res 2003;31:e61.

9. St John JC. Mitochondrial DNA copy number and replication in reprogramming and differentiation. Semin Cell Dev Biol 2016;52:93–101.

10. Alberts B, Johnson A, Lewis J, Raff M, Roberts K, Walter P. Molecular Biology of the Cell. New York: Garland Publishing Inc. ISBN 0-8153-3218-1. 1994.

11. Moyes CD, Battersby BJ, Leary SC. Regulation of muscle mitochondrial design. J Exp Biol 1998;201:299–307.

12. Mengel-From J, Thinggaard M, Dalgard C, Kyvik KO, Christensen K, Christiansen L. Mitochondrial DNA copy number in peripheral blood cells declines with age and is associated with general health among elderly. Hum Genet 2014;133:1149–59.

13. Ding J, Sidore C, Butler TJ, et al. Assessing Mitochondrial DNA Variation and Copy Number in Lymphocytes of ~2,000 Sardinians Using Tailored Sequencing Analysis Tools. PLoS Genet 2015;ll:el005306.

14. Zhang R, Wang Y, Ye K, Picard M, Gu Z. Independent impacts of aging on mitochondrial DNA quantity and quality in humans. BMC Genomics 2017;18:890.

15. Xu FX, Zhou X, Shen F, Pang R, Liu SM. Decreased peripheral blood mitochondrial DNA content is related to HbA1c, fasting plasma glucose level and age of onset in type 2 diabetes mellitus. Diabet Med 2012;29:e47-54.

16. Lee JY, Lee DC, Im JA, Lee JW. Mitochondrial DNA copy number in peripheral blood is independently associated with visceral fat accumulation in healthy young adults. Int J Endocrinol 2014;2014:586017.

17. Guyatt AL, Burrows K, Guthrie PAI, et al. Cardiometabolic phenotypes and mitochondrial DNA copy number in two cohorts of UK women. Mitochondrion 2018;39:9–19.

18. Tin A, Grams ME, Ashar FN, et al. Association between Mitochondrial DNA Copy Number in Peripheral Blood and Incident CKD in the Atherosclerosis Risk in Communities Study. J Am Soc Nephrol 2016;27:2467–73.

19. Scott RA, Lagou V, Welch RP, et al. Large-scale association analyses identify new loci influencing glycemic traits and provide insight into the underlying biological pathways. Nature genetics 2012;44:991–1005.

20. Levy D, Ehret GB, Rice K, et al. Genome-wide association study of blood pressure and hypertension. Nat Genet 2009;41:677–87.

21. Wilier CJ, Schmidt EM, Sengupta S, et al. Discovery and refinement of loci associated with lipid levels. Nat Genet 2013;45:1274–83.

22. Huang PL. A comprehensive definition for metabolic syndrome. Dis Model Mech 2009;2:231–7.

23. WHO. Waist Circumference and Waist–Hip Ratio: Report of a WHO Expert Consultation Geneva, 8–11 December 2008 2008.

24. Ashar FN, Moes A, Moore AZ, et al. Association of mitochondrial DNA levels with frailty and all-cause mortality. J Mol Med (Berl) 2015;93:177–86.

25. Houseman EA, Accomando WP, Koestler DC, et al. DNA methylation arrays as surrogate measures of cell mixture distribution. BMC Bioinformatics 2012;13:86.

26. Abdi H. Partial least squares regression and projection on latent structure regression (PLS Regression). WIREs Computational Statistics 2010;2:97–106.

27. Liu C, Dupuis J, Larson MG, Levy D. Association testing of the mitochondrial genome using pedigree data. Genetic epidemiology 2013;37:239–47.

28. Lee HK, Song JH, Shin CS, et al. Decreased mitochondrial DNA content in peripheral blood precedes the development of non-insulin-dependent diabetes mellitus. Diabetes Res Clin Pract 1998;42:161–7.

29. Vozarova B, Weyer C, Lindsay RS, Pratley RE, Bogardus C, Tataranni PA. High White Blood Cell Count Is Associated With a Worsening of Insulin Sensitivity and Predicts the Development of Type 2 Diabetes. Diabetes 2002;51:455–61.

30. Dixon JB, O’Brien PE. Obesity and the white blood cell count: changes with sustained weight loss. Obes Surg 2006;16:251–7.

31. Kullo IJ, Hensrud DD, Allison TG. Comparison of numbers of circulating blood monocytes in men grouped by body mass index (<25, 25 to <30, > or =30). Am J Cardiol 2002;89:1441–3.

32. Ohshita K, Yamane K, Hanafusa M, et al. Elevated white blood cell count in subjects with impaired glucose tolerance. Diabetes Care 2004;27:491–6.

33. Twig G, Afek A, Shamiss A, et al. White blood cells count and incidence of type 2 diabetes in young men. Diabetes Care 2013;36:276–82.

34. Orakzai RH, Orakzai SH, Nasir K, et al. Association of white blood cell count with systolic blood pressure within the normotensive range. Journal of Human Hypertension 2006;20:341–7.

35. Karthikeyan VJ, Lip GYH. White blood cell count and hypertension. Journal of Human Hypertension 2006;20:310–2.

36. Gillum RF, Mussolino ME. White blood cell count and hypertension incidence. The NHANES I Epidemiologic Follow-up Study. J Clin Epidemiol 1994;47:911–9.

37. Ashar FN, Zhang Y, Longchamps RJ, et al. Association of Mitochondrial DNA Copy Number With Cardiovascular Disease. JAMA Cardiol 2017;2:1247–55.

38. Bellizzi D, D’Aquila P, Giordano M, Montesanto A, Passarino G. Global DNA methylation levels are modulated by mitochondrial DNA variants. Epigenomics 2012;4:17–27.

39. Latorre-Pellicer A, Moreno-Loshuertos R, Lechuga-Vieco AV, et al. Mitochondrial and nuclear DNA matching shapes metabolism and healthy ageing. Nature 2016;535:561–5.

40. Castellani CA, Longchamps RJ, Sumpter JA, et al. Mitochondrial DNA Copy Number (mtDNA-CN) Can Influence Mortality and Cardiovascular Disease via Methylation of Nuclear DNA CpGs. bioRxiv 2019:673293.

41. Lai YC, Woollard KJ, McClelland RL, et al. The association of plasma lipids with white blood cell counts: Results from the Multi-Ethnic Study of Atherosclerosis. J Clin Lipidol 2019;13:812–20.

42. Wu IC, Lin CC, Liu CS, Hsu CC, Chen CY, Hsiung CA. Interrelations Between Mitochondrial DNA Copy Number and Inflammation in Older Adults. J Gerontol A Biol Sci Med Sci 2017;72:937–44.

43. Knez J, Marrachelli VG, Cauwenberghs N, et al. Peripheral blood mitochondrial DNA content in relation to circulating metabolites and inflammatory markers: A population study. PLoS One 2017;12:e0181036.

44. Sharma P. Inflammation and the metabolic syndrome. Indian J Clin Biochem 2011;26:317–8.

45. Esposito K, Giugliano D. The metabolic syndrome and inflammation: association or causation? Nutr Metab Cardiovasc Dis 2004;14:228–32.

46. Saltiel AR, Olefsky JM. Inflammatory mechanisms linking obesity and metabolic disease. J Clin Invest 2017;127:1–4.

47. Paoletti R, Bolego C, Poli A, Cignarella A. Metabolic syndrome, inflammation and atherosclerosis. Vase Health Risk Manag 2006;2:145–52.

48. Short KR, Bigelow ML, Kahl J, et al. Decline in skeletal muscle mitochondrial function with aging in humans. Proc Natl Acad Sci U S A 2005;102:5618–23.

49. Kaaman M, Sparks LM, van Harmelen V, et al. Strong association between mitochondrial DNA copy number and lipogenesis in human white adipose tissue. Diabetologia 2007;50:2526–33.

50. Xing J, Chen M, Wood CG, et al. Mitochondrial DNA content: its genetic heritability and association with renal cell carcinoma. J Natl Cancer Inst 2008;100:1104–12.

51. Yang SY, Castellani CA, Longchamps RJ, et al. Blood-derived mitochondrial DNA copy number is associated with gene expression across multiple tissues and is predictive for incident neurodegenerative disease. bioRxiv 2020:2020.07.17.209023.

